# Nursing Intervention Analysis in COVID-19 Negative Pressure Isolation Wards and General Wards: Observational study

**DOI:** 10.1101/2023.01.06.23284223

**Authors:** Hyunsoon Park, Mi sug Lee, Lee Gyu Min, Hee Oh, Sung Hwangbo, Sanghyuk Roh, Ho Heon Kim

**Affiliations:** Department of nursing, Gumi University, Gyeongsangbuk-do, Republic of Korea; Department of nursing, Bestian Hospital, Chungcheongbuk-do, Republic of Korea; Department of nursing, Chung Goo Sung Sim Hospital, Seoul, Republic of Korea; Department of nursing, Ansan University, Gyeongsangbuk-do, Republic of Korea; Department of nursing student, Gumi University, Gyeongsangbuk-do, Republic of Korea; Department of Biomedical Systems Informatics, College of Medicine, Yonsei university, Seoul, Republic of Korea

**Keywords:** Health provider burden, Nursing management, Negative pressure ward, Time Motion study, Nursing Intervention

## Abstract

**Introduction:** With the global spread of Coronavirus disease (COVID-19) and public health crisis, appropriate allocation of healthcare human resources has been necessitated. Although nursing practice takes up a larger part of medical practice in hospitals, the quantitative assessment of nursing care has not been investigated for human resource allocation in the medical field. The objective of this study to explore the time spent for each nursing intervention, and compared provided amount of nursing intervention between negative pressure isolation wards (NPIWs) and general wards (GWs) provided by COVID-19 hub hospitals.

**Methods:** This research is a time-motion (TM) observational study. Three trained external observers recorded their observations for every minute in 19 different work schedules in 2 NPIWs and 2 general respiratory wards. Observation items were chosen based on the standard operating guidelines of Integrated Nursing and Caring Services developed by the Ministry of Health and Welfare and National Health Insurance Service. The average nursing workload per shift was compared by calculating the sum of the spent time of three nurses staffed in each shift in each ward between two groups. In addition, to compare the amount of directed nursing care for patients between two types of wards, nursing work category was divided into directed and undirected nursing interventions.

**Results:** In the comparison of demographic characteristics of nursing workforce between two groups, there was no statistically significant difference (p>0.05 respectively). In both groups, the most time-consuming nursing work category was recording in three work shifts. The average duration of those work tasks was 312.5 minutes in NPIWs and 307 minutes (per 3 nurses) in GWs, having no significant difference (p>0.05). Of all nurse duties, the second most time-consuming work category was others (including changing to protective clothing) in NPIWs, and medication administration and transfusion in GWs. The mean duration of performing the category for others that include wearing PPE was 308 minutes in NPIWs and 160 minutes (per 3 nurses) in GWs, showing a significant difference (p<0.05). The greater amount of time was taken for hygiene management in isolation wards. Medication administration and transfusion and nursing assessment were more frequently performed in GWs, demonstrating a statistical significance. In the aggregated spent time for all duties including directed and undirected nursing care, the time spent for directed nursing care was 654 minutes longer in GWs than in NPIWs (per 3 nurses) in each work shift, displaying a significant difference.

**Conclusion:** This study provides the quantitative difference in time-consuming nursing works between NPIWs and GWs by direct observation. Recording was the most time-consuming nursing work category in both NPIWs and GWs. Considering nurses in each duty in GWs provided more directed nursing care than nurse in NIPWs, careful considerations are required in allocation of nursing workforce.

## Introduction

The pandemic of Coronavirus disease 2019 (COVID-19) has brought substantially changes in duties in the nursing field. The WHO first reported COVID-19 on December 31, 2019 [WHO. 2003], and declared Public Health Emergency of International Concern (PHEIC) international attention on January 31, 2020 [Kim and Lee, 2021]. Since the first outbreak of severe acute respiratory syndrome (SARS) in 2002 in China, disease spread has influenced not only SARS affected patients, but also healthcare providers in wards and regions. The repeated experiences of infectious disease pandemic disasters have provided an opportunity to warn medical negligence on infectious disease and reform prevention measures worldwide for the past decades.

Unlike other diseases, infectious diseases are very extensive and continuously use up medical resources. In particular, healthcare providers, the most important medical resources have been consistently consumed by patients with infectious disease. Recent studies have revealed that the outbreak of COVID-19, one of the infectious diseases has threaten mental health and lives of nurses by increasing tension and stress in nursing duties [Kackin, 2021]. Middle East respiratory syndrome coronavirus (MERS-CoV), one of recent infectious diseases has caused the risk of post-traumatic stress disorder (PTSD) for a substantial period of time in healthcare workers who participated in clinical management of this pandemic. Likewise, infectious disease crisis can incur negative outcomes short- and long-term in both physical and mental health of medical staffs. Therefore, research for prevention and management of infectious disease needs to be prepared for public health in a broad perspective as we go through the era of coexisting with coronavirus disease.

In practice, intensive care units (ICUs) are being operated for crisis response to a surge of confirmed COVID-19 cases in the clinical fields. For prevention of airborne transmission of COVID-19, confirmed patients are admitted to negative pressure isolation wards (NPIWs) with the control system of infectious aerosols escape, instead of in general wards (GWs) [Eom, 2018]. Nurses working in NPIWs have additional tasks compared to those working in GWs. These tasks included work environment of exposure to infectious disease, work stress in isolation wards (IWs), wearing full-body protective clothing as personal protective equipment (PPE), attachment of Easy Talk device for two-way communication, wearing a powered air purifying respirator (PAPR) and others.

With the worldwide spread of the COVID-19 pandemic, adequate allocation of medical resources is demanded for healthcare providers in the crisis of public health [Lee, 2020]. Even though nursing intervention accounts for one of the largest proportions of medical practices, nursing care has been insufficiently considered and explored in allocation of healthcare human resources and quantitative assessment of nursing intervention. Therefore, understanding nursing workload and the time spent on each nursing intervention in NPIWs is required to optimal nurse staffing [Kalisch et al., 2011], [Ren et al., 2022], and achieve patient outcomes [Yi & Kim, 2021; Xie, 2021]. However, studies have not yet been performed to explore the total duration of nursing intervention and total nursing workload. In addition, no studies have compared NPIWs and GWs by observing the subjects on the basis of time-based patterns. This research designed as a time-motion (TM) observational study aims to explore the time spent for each nursing intervention, and compared provided amount of nursing intervention between negative pressure isolation wards (NPIWs) and general wards (GWs) provided by COVID-19 hub hospitals

## Method

### Overall Study Design

In this cross-sectionally observational study, we recruited the four secondary general hospitals that have more than 100 beds and less than 300 beds. Among four hospital, two hospitals with isolation wards for COVID-19 which designated by Ministry of Health and Welfare were participated to figure out the overall work hours for each activity of a nurse, and difference in duration of directed/undirected nursing cares. The nursing delivery system of two isolation wards is functional nursing (task-forced), however, that of general wards is team nursing (person-centered). In addition, the all of duties in each ward are staffed by three nurses. Therefore, we compared the total duration of each nursing work category by shift (of three nurses) in isolation wards and in general wards to consider the different nursing delivery system. To assess the time required to accomplish specific nurses’ tasks, we conducted basic time motion study (TMS) [Taylor, 1914; Lopetegui, 2014].

### Participants Recruitment

Four hospitals were selected as study samples through convenience sampling including two COVID-19 hub hospitals with isolation wards and two hospitals with general wards located in Seoul, Gyeonggi, and Chungbuk regions among 318 tertiary hospitals registered to the Health Insurance Review and Assessment Service as healthcare institutions in 2022 [Korean Statistical Information Service (KOSIS), 2022]. This study recruited samples after gaining the approval of participation from each institution. One of the methods for efficient use of nursing workforce is to utilize the patient classification system. This study classified patient severity in each hospital using the Korean Patient Classification System (KPCS-1).

The subjects were chosen through population sampling by posting a recruit notice and gaining approval in the Department of Nursing of each hospital. To avoid any negative consequences by co-investigators in subject selection, exogenous variables were controlled with recruitment of study subjects. All subjects were fully informed of the research purposes and survey procedures and voluntarily participated to the study with a written consent among formally employed nurses working in NPIWs and GWs. Inclusion criteria for performance ability of nursing interventions were a minimum clinical experience of two years on average, work position of charge nurse with work experience in NPIWs, and a minimum educational level of bachelor degree in nursing. After the classification of patient severity by the Department of Infection in each hospital chosen as the participating institution, patient information retrieval was extracted by nurses who were the third persons among confirmed COVID-19 patients admitted to NPIWs.

### Questionnaire items

This study investigated the clinical practice guidelines of international, domestic and foreign, and hospital standards to build observational questionnaire items for nursing interventions in COVID-19 patients in NPIWs. We selected candidate observational items as following: 1) A literature review and information retrieval on domestic and international clinical practice guidelines in NPIW nursing intervention; 2) Detailed work tasks according the clinical nursing practice guideline categories were divided into 101 items in 13 different categories chosen based on the standard operating guidelines of Integrated Nursing and Caring Services developed by the Ministry of Health and Welfare and National Health Insurance Service [the Ministry of Health and Welfare; National Health Insurance Service, 2016] and the response measures of government organizations and local communities according to COVID-19 crisis response situations of the Korea Research Institute for Healthcare Policy [KHAPA, 2021]. 3) Job description, clinical nursing manual, and clinical practice guidelines equipped in each ward were analyzed to identify work tasks and work flow patterns in NPIWs. 4) Based on the above mentioned stages, nursing intervention items addressed in COVID-19 NPIW clinical nursing practice guidelines were developed as the directed and undirected nursing intervention tool consisting of 19 categories and 123 items. To ensure the reliability of the developed survey tool, discussed issues in NPIWs were reflected through simulation prior to the main study. Comments and opinions from expert nurses were shared and member check were additionally done to improve the validity of the study results [Birt et al., 2016].

For the assessment of directed and undirected nursing interventions as the COVID-19 NPIW clinical nursing practice guidelines, the study tool developed in this study consists of 19 work categories and 123 items on detailed work tasks. The 19 major nursing practice categories include respiration (10), medication and transfusion (5), nutrition (3), safety (8), measurement and observation (13), excretion (13), hygiene (9), exercise and activity (10), stability (8), treatment and inspection (7), information management (4), check-up and follow-up (7), communication (6), nurse training (1), education and consultation (7), emotional support (3), communication/adjustment between departments (3), personal free time, and others (5).

### Data collection

Data were collected after gaining the approval from the Department of Nursing of each institution participating the study. Data collection for the NPIW nursing intervention assessment tool was carried out until November after gaining the IRB approval. Observation was done by making a contract to the chief director of each hospital prior to the visit. As protective measures in the direct observation of vulnerable subjects, to exclude any concerns over possible negative consequences by co-investigators who could affect subject’s decision-making for voluntary participation to the study, consent was obtained by the third person unrelated to this study.

Data collection was performed by external observers. Prior to the direct observation of the subjects, the participants were fully informed of the research purposes and procedures by the observer in every work shift. Investigators saved basic data in the Stopwatch a tablet computer, and recorded direct observation checkpoints and took note of directed and undirected nursing interventions in COVID-19 patients.

The time spent on nursing interventions were measured in details according to the flow of work from the starting point to the ending point in every minute. The spent time was analyzed by examining only the floor where the nurse station was located considering spatial location and facility arrangement. The analysis was performed by selecting hospital rooms having the similar distance from the nurse station and patient room. For 24-hour straight data collection as a TM observational study, the principal and co-investigators visited four hospitals and consecutively observed 36 nurses working on their day, evening, and night shifts by nursing intervention without rest. Not to disturb work during observation time, observers exerted effort to perform 1:1 observation at a close distance from 7 a.m. to 7 a.m. the next day. Nursing interventions done for patients in NPIWs and GWs were explored in details including the content, frequency, and nursing care hours of interventions.

### Statistical analysis

Descriptive statistics for nurses’ profile (eg, age, education, position) and work profile including working hours, were explored. Then, we conducted Fisher’s exact test to determine the difference in the proportion of categorical variables of nurses working categories in isolation wards, and general wards by shift by considering small-sized samples[Kim, 2017]. For continuous variables such as work hours recorded of each nursing category in seconds, Mann Whitney U-test was performed. We consider *P* values <0.05 as statistical significance for 2-sided hypothetical tests. By aggregating work categories according to the classification between the directed or undirected nursing cares, we analyzed the difference in the total time of directed or undirected nursing care between isolation wards and general wards. We conducted statistical analysis with python (version 3.7) including open source package Table One, and Pandas [Tom J Pollard, 2018].

### Ethics Statement

The study protocol was approved by the Institutional Review Board at Advanced Clinical Trial Center of Bestian Hospital (IRB No. 2022-02-003-004). We directly obtained voluntary informed consent for participation in the research by explaining the research purpose and method. This study was supported by supported by Small and Medium Hospital Nursing Association and Gyeonggi-do Nurses Association in 2022 (No. 202206-14).

## Result

### Characteristics of Participants

In total 36 of participants, each 18 of participants were enrolled in both isolation ward, and general wards. The median of age in both groups did not show statistically significant difference (p=0.975). Also, there is no statistical significance in proportion of education, and marital status position. However, work experience of nurses in general wards significantly higher than those of nurse in isolation ward (p<0.037). In addition, their median of extended work hours did not show significantly difference (p=0.071) (Table 1).

**Table 1.** Demographic characteristics of participants

| Variables |  | Participants<br>(N=36) |  | P-value |
| --- | --- | --- | --- | --- |
|  |  | Isolation wards<br>(n=18) | general wards<br>(n=18) |  |
| Age, (median [Q1,Q3]) |  | 28.5 [26.0,42.5] | 29.0 [26.2,33.8] | 0.975 |
| Marital Status, (n, (%)) | Married | 5 (27.8) | 3 (16.7) | 0.691 |
|  | Not married | 13 (72.2) | 15 (83.3) |  |
| Education, (n, (%)) | Associated degree | 6 (33.3) | 1 (5.6) | 0.088 |
|  | Bachelor | 12 (66.7) | 17 (94.4) |  |
| Position, (n, (%)) | Head nurse | 2 (11.1) |  | 0.346 |
|  | Charge nurse | 1 (5.6) | 1 (5.6) |  |
|  | Nurse | 15 (83.3) | 17 (94.4) |  |
| Number of patients per nurse,<br>(n(nurse), (%)) | Over 14 | 18 (100.0) | 4 (22.2) | <0.001 |
|  | 12 | 0 (0.0) | 10 (55.6) |  |
|  | 10 | 0 (0.0) | 4 (22.2) |  |
| Work experience, (years,<br>median [Q1,Q3]) | In current ward | 9.0 [3.0,12.0] | 34.5 [6.0,40.5] | 0.037 |
|  | Total work experience | 60.0<br>[24.0,165.0] | 60.0 [39.0,96.0] | 0.874 |
| Work type (n, (%)) | Three shifts | 18 (100.0) | 16 (88.9) | 0.486 |
|  | Fixed night shift | 0 (0.0) | 2 (11.1) |  |
| Nursing delivery system (n,<br>(%)) | Functional | 18 (100.0) | 0 (0.0) | <0.001 |
|  | Team | 0 (0.0) | 18 (100.0) |  |
| Extended work hours (n, (%)) | > 4 hours | 4 (22.2) | 0 (0.0) | 0.071 |
|  | < 4 hours and 1 > hours | 14 (77.8) | 17 (94.4) |  |
|  | None | 0 (0.0) | 1 (5.6) |  |

### Overall spent time for each nursing work category by shifts

Figure 1 showed the overall spent time for each nursing work category aggregated by shift. In all of duties in each ward, recoding time was the largest proportion of working hours. On averaged considering all shifts in all wards, 309.25 of minutes was spent to record ENRs by shift (SD: 53.9). For general wards, the spent time to conduct medication and transfusion was the secondary largest proportion, on the other hand, nurses in isolation ward spent time working on task classified as others. On day shift in isolation wards, 188, and 124 of communication time was observed, which was larger than that of evening shift (82.0, 71.0; respectively) and night shift (108.0, 69.0; respectively).

**Figure 1.**
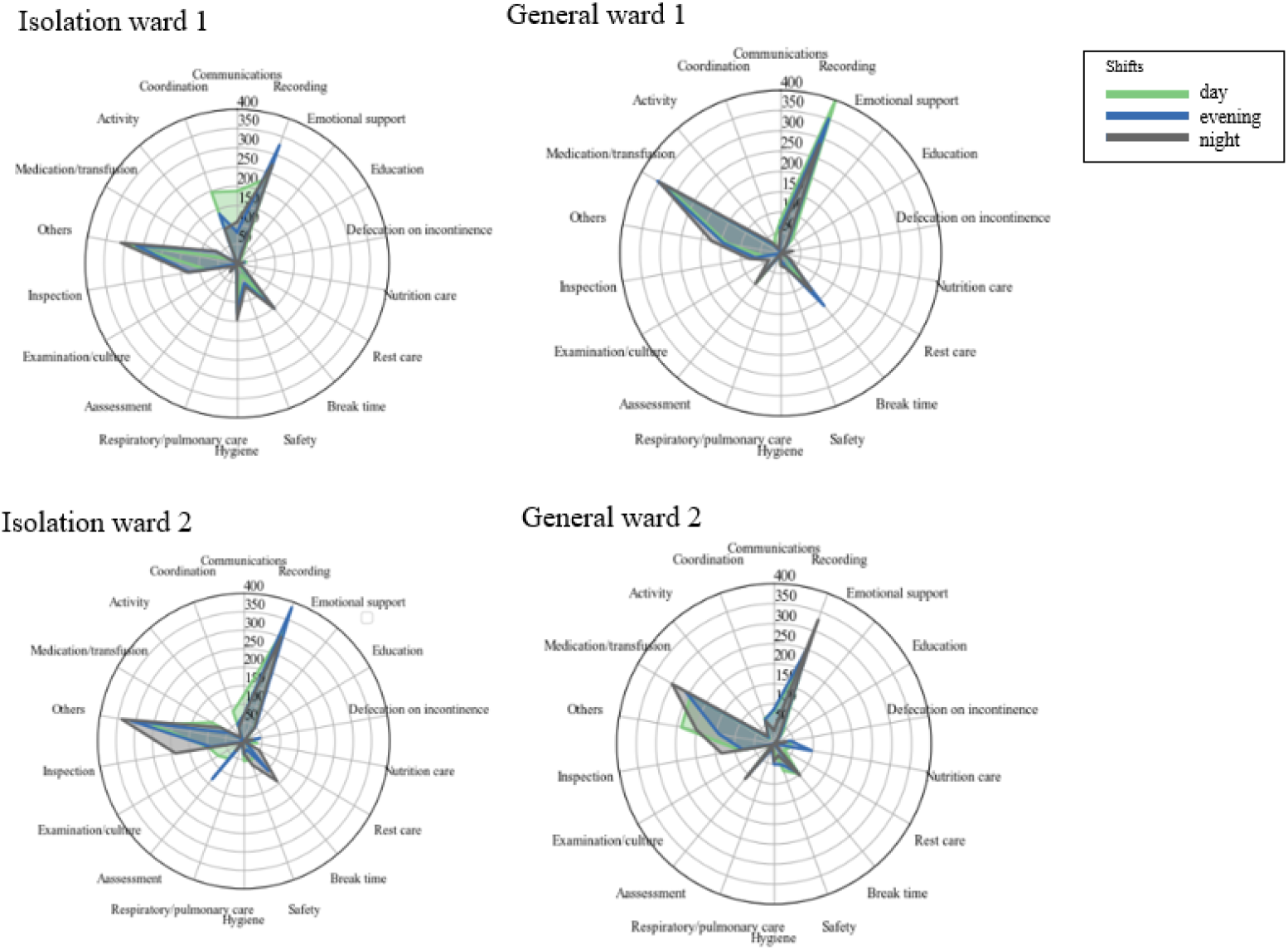
Overall spent time for each nursing work category aggregated by shift. For each shift, three nurses were staffed in each ward. Therefore, the total amount of spent time for all work categories is 1440(8 hours * 60 minutes * 3 nurses).

### Categories of nursing in working hours

Table 2 showed that the sum of the spent time by duties, which were staffed by three nurses, to perform each nursing work category. The most time-consuming nursing working category in isolation ward was recording, followed by others, breaktime, inspection management. On the other hand, in general wards, they spent most of time to conduct medication and transfusion, followed by recording, and other, breaktime, assessment. In addition, median of aggregated spent time by duties to perform the other, hygiene management, and education showed significant difference (p<0.05, respectively, Table 2).

**Table 2.** Comparison of work categories between isolation and general wards

| Nursing work categories<br>(sum of minutes by duties, (median, [Q1, Q3])) | Total duties<br>(N=12) |  | P-value |
| --- | --- | --- | --- |
|  | duties<br>in isolation wards<br>(n=6) | duties<br>in general wards<br>(n=6) |  |
| **Recording** | **312.5 [280.0,326.2]** | **307.0 [272.2,345.5]** | **0.873** |
| **Others** | **308.0 [278.8,312.5]** | **160.0 [142.8,193.8]** | **0.004** |
| Break time | 145.0 [115.0,151.0] | 100.5 [94.2,114.2] | 0.337 |
| **Inspection management** | **112.5 [92.8,130.8]** | **80.0 [68.5,87.0]** | **0.055** |
| General communications | 95.0 [73.8,120.0] | 72.0 [55.5,81.0] | 0.078 |
| Coordination with other health providers | 90.0 [58.0,126.5] | 51.0 [18.5,64.0] | 0.078 |
| **Hygiene management** | **87.0 [41.2,127.5]** | **30.0 [20.2,40.5]** | **0.024** |
| **Medication and transfusion** | **64.5 [49.0,71.8]** | **314.0 [269.0,343.2]** | **0.004** |
| Safety management | 55.0 [36.8,62.0] | 37.0 [32.0,51.0] | 0.688 |
| Emotional support | 30.5 [9.8,44.5] | 24.0 [20.8,31.0] | 1.000 |
| **Assessment** | **30.5 [26.5,54.8]** | **93.0 [84.8,96.0]** | **0.078** |
| Nursing for defecation on incontinence | 24.0 [21.0,24.0] | 31.0 [24.2,37.8] | 0.518 |
| Education | 23.5 [10.5,29.0] | 52.0 [44.0,63.0] | 0.149 |
| Nursing for rest | 9.5 [3.5,17.8] | 15.0 [7.2,25.0] | 0.630 |
| Examination and culture | 8.5 [4.2,13.5] | 3.5 [2.0,5.0] | 0.147 |
| Respiratory and pulmonary nursing care | 4.5 [3.2,6.5] | 7.5 [5.5,8.0] | 0.257 |
| Nursing for activity | 3.5 [0.8,4.8] | 24.0 [20.8,25.0] | 0.004 |
| Nursing for nutrition | 1.5 [0.0,8.2] | 4.0 [3.0,35.0] | 0.286 |

We additionally analyzed the work categories in others in Figure 2. Comparison of subcategorized nursing in “other” categories between isolation and general wards. Not significant was denoted by ns. Statistical significance in difference in median for each value denotes was denoted by *, and ** (p<0.05, p<0.01). showed difference in median value of work hours for other categories work by nurses in both wards. Two subcategories of works such as wearing PPE or medical equipment, and other communication showed significant difference in median value of it (p<0.05, respectively, Figure 2).

**Figure 2.**
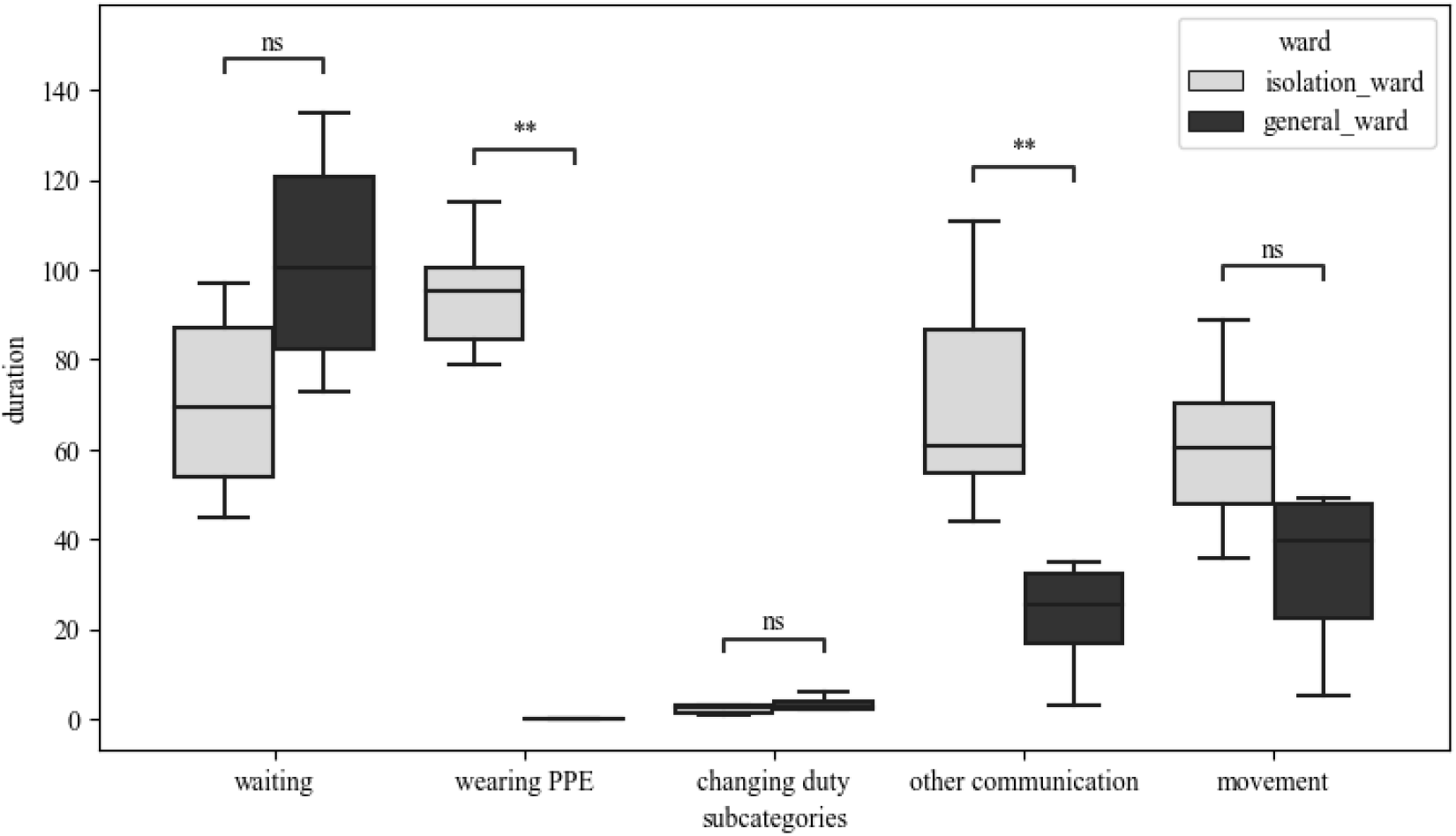
Comparison of subcategorized nursing in “other” categories between isolation and general wards. Not significant was denoted by ns. Statistical significance in difference in median for each value denotes was denoted by *, and ** (p<0.05, p<0.01).

### Directed and Undirected nursing in each wards

For isolation wards, and general wards, the average of 379.8, and 654 minutes for directed nursing care by duties were observed respectively, Also, their average time for directed nursing care showed significant difference in both groups (p<0.005) (Figure 3).

**Figure 3.**
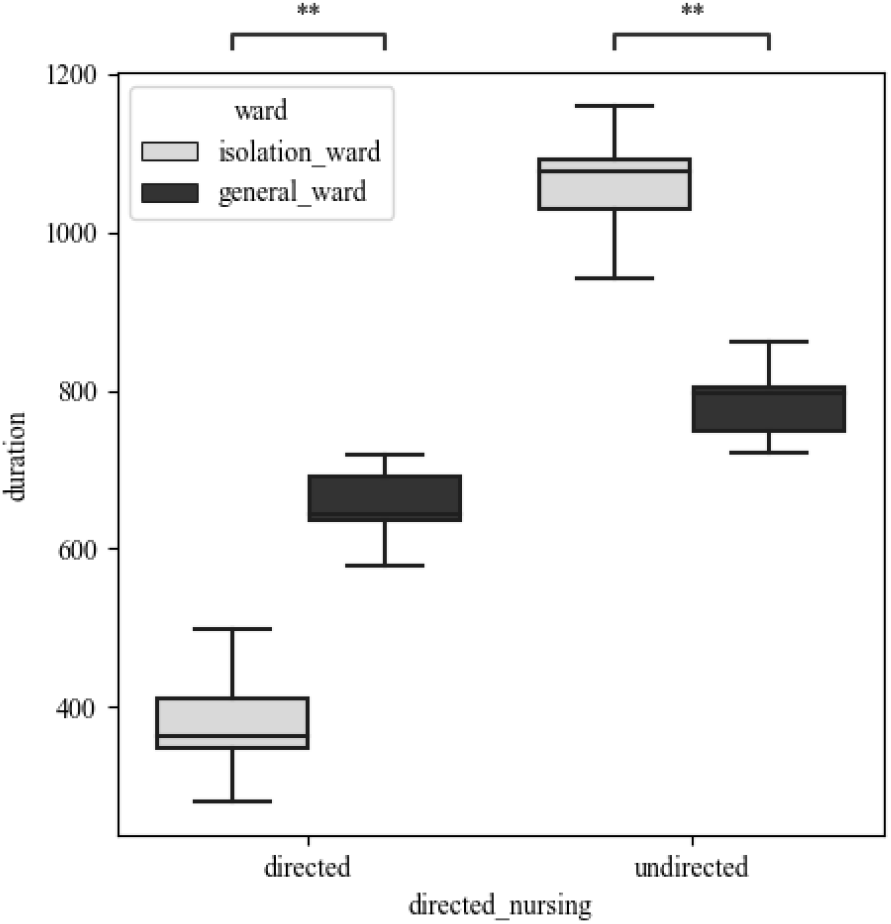
Overall directed and undirected nursing in each shift between isolation and general wards by duties

## Discussion

### Principal findings

As consumption of medical resources has been concentrated in healthcare workforce due to COVID-19 spread, strategies for optimal allocation of scarce medical resources have been demanded more than ever before. Previous studies focused on gross measurements, for example, the number of beds in the emergency department, the number of beds in a NPIW were mainly focused on [Emanuel, 2020]. However, to optimize medical resource allocation, this study quantitatively assessed whether what kind of and how much nursing duties were performed in NPIWs and GWs. Recently, the supply of medical resources should be distributed according to nursing interventions, and the need of scientific evidence has gained much attention for adequate and fair allocation. This basic research study on the scientific evidence observed nurses’ duties and workload in both NPIWs dedicated to infectious disease and general respiratory wards and found out the difference in work hours according to nursing duties between IWs and GWs.

### Optimization of nurse allocation for isolation wards

For each shift in NPIWs, nurses took additional burden related to changes to PPE, and communication were additional (Figure 1). The median value of time spent for these works was 308 minutes for three nurses, approximately 1.92 times higher than that of GW. The time spent on this work category accounted for 20% (308/1,440 minutes=3 nurses*8 hrs*60 mins) of all working hours, taking a large proportion in each shift of three nurses (Table 2). This difference seems to be resulted from the more time spent on undirected care using SNS and mobile phones for communication in NPIWs. Instead of direct observation for nursing assessment, monitoring outcomes were self-reported by patients via text messages or phone calls. Moreover, the longer duration of wearing PPE was identified as the difference in disease control and management methods in NPIWs than in GWs (Figure 2).

In the comparison of the total time spent on directed and undirected nursing cares, fewer hours were spent on directed care for patients admitted to NPIWs (Figure 3). The duration of directed nursing care was 379 minutes per shift among NPIW nurses, spending 0.579 times lesser than 654 minutes per shift among GW nurses. The difference in directed and undirected cares is thought to be attributable to the fact that undirected nursing care is more needed for NPIW nurses in the following categories including wearing PPE, communication, moving, and inspection management. When the same amount of directed care is required for the same patient, this difference implies that 1.72 nurses, the inverse value from numerical calculation are needed to meet the requirement of directed nursing care. Considering that the number of medical staffs has a negative correlation with patient’s outcome, this should be taken into consideration in calculating nurse staffing when the same directed care is needed [Xie, 2021]. To ensure the same amount of nursing care, a buffer in needed to preserve nurse staffing calculated in the above [Yin, 2021].

### To mitigate burden of nursing staff working for negative pressure isolation wards

During the COVID-19 pandemic, nursing workforce has been proven to be an indispensable resource in the health system [Llop-Gironés, 2021]. The continuous spread of COVID-19 has aggravated inequalities between demand and supply for nurses as nurses have experienced more burnout. Several studies commonly suggested “the balance between patient and nurse needs satisfaction and exhaustion management” to help ease nurses’ burnout experience during the COVID-19 pandemic [Roe, 2022]. In particular, excessive work duties and wearing nearly 3kg-protective equipment were identified to be related to increased fatigue [Liu Q, 2020; Iheduru-Anderson, 2021]. In line with the results of a preliminary study, the findings of this study also demonstrated that PPE, communication and others had a significant impact on nursing workload.

First, needs for nursing care can be satisfied by reducing the amount of nursing care or increasing the supply of nursing workforce. Although the public healthcare sector has exerted a range of efforts to decrease the high demand of nursing care with multiple preventive strategies, more studies and policies are warranted to increase the supply of nursing staff. However, to the best of our knowledge, no studies have not yet quantitatively assessed nurse staffing by ward. This review observed that there was a difference in the amount of duties performed by nurses according to their work shifts (Figure 1). In particular, time spent on intervention and communication among healthcare providers increased during the day shift in NPIW. This outcome is anticipated to be resulted from frequent physician rounds in the day duty, and major nursing duty schedules are mostly done during the day time. For nurses specialized in these kinds of tasks, undirected nursing care can be shifted to direct nursing care. Moreover, additional studies are warranted to further investigate time spent for wearing PPE, two-way communication devices and others.

Second, moving line optimization, development of highly usable PPE and others can be taken into consideration in fatigue management for nurses. As effective measures to reduce fatigue for nurses, moving line optimization is a critical factor in the increase of directed nursing care. Unlike in general wards, the environmental structure of passing through entry and exit doors twice for assess to isolation rooms, in particular, seems to increase undirected nursing care time. To design NPIWs more efficiently, the mandatory installation of clean rooms [Cho and Seong, 2018] and pass boxes [National Medical Center, 2021] is expected to contribute to reducing undirected nursing care time. Placement of the pass box equipped with an ultraviolet sterilizing device between the nurse station and hallway of the NPIW is thought to reduce undirected time spent for PPE wearing in the absence of nursing and inspection supplies missing in two-way communication. The common physical problems associated with using PPE kits were excessive sweating (100%), fogging of goggles, spectacles, or face shields (88%), suffocation (83%), breathlessness (61%), fatigue (75%), headache due to prolonged use (28%), and pressure marks on the skin at one or more areas on repeated use (19%) [Agarwal, 2020] These problems need to be further clarified by performing additional studies. Moreover, as shown in the study findings, considering the time required taking about 20% of the total working hours as a barrier to the use of PPE, reduced duration of wearing PPE and light weight of PPE kits will be helpful in considerably reducing burden toward directed nursing care for long periods of time. Barriers to using PPE reported were inappropriate PPE size, design of PPE and complexity of use, doubts related to the quality and effectiveness of PPE, potential risks during doffing, and others [Fan, 2020]. The use of sterile gloves and disposable shoe covers in medical practices needs to be included in the future studies. Overcoming the above barriers is forecasted to help manage fatigue in nurses.

More investigations are crucial to standardize guidelines based on the time spent on nursing interventions to improve care quality associated with reduced missing rate of interventions and calculate nurse staffing in acute infectious disease control. A systematic study is warranted to further elucidate the association between consideration on effective environmental structure and reduction of the total spent time by duties.

## Limitations

This study was limited in certain aspects. First, the research findings on hospitals voluntarily participated in this survey have limitations to be generalized to represent all COVID-19 hub hospitals and general hospitals. However, this study was meaningful in that it contributed to ensuring reliability with an effort to demonstrate simulation prior to the main observational survey considering the work environment of COVID-19 NPIW and selection of nursing intervention boundaries of nursing practice with the development of a directed and undirected nursing recording form. Second, 24-hour observation of three co-investigators unrelated to vulnerable research participants of this study appears to help overcome research bias. The findings of this study are meaningful in that it intended to define the scope and boundaries of NPIW nursing intervention in clinical practices and to assess nurse staffing for reduction of missing rate of interventions in acute infectious disease control.

## Conclusion

This study provides the evidence that the differences in nursing work pattern between NPIWs and GWs, and nurses in NPIWs spent more time on undirected care. To ensure the equal level of nursing care in NPIWs with GW, a extra nurse workforce may be needed. In addition, efforts to reduce the burden on nursing personnel working at NPIW should be considered.

## Data Availability

Not available

## Abbreviation

COVID-19: Coronavirus Disease 2019
GW: General Wards
IRB: Institutional Review Board
NPIW: Negative Pressure Isolation Wards
SD: Standard Deviation

